# Burr-Hole Intersection of Middle Meningeal Artery Branches and Recurrence in Chronic Subdural Haematoma: a Multicentre Retrospective Cohort Study

**DOI:** 10.64898/2026.08.26.26361348

**Authors:** Tymour M Saba, Jigi Moudgil-Joshi, Anand S Pandit, Jack Penn, Dermot Mallon, Hani J Marcus, Patrick Grover

## Abstract

**Background and Objectives:** Recurrence following burr-hole drainage of chronic subdural haematoma (cSDH) occurs in 10-25% of cases, sustained by neovascularisation of the subdural neomembrane supplied by the middle meningeal artery (MMA). MMA embolisation reduces recurrence; whether incidental burr-hole intersection of MMA branches during drainage confers similar benefit is unknown.

**Methods:** We performed a multicentre retrospective cohort study of consecutive adults undergoing burr-hole drainage for cSDH at two UK tertiary neurosurgical centres. Postoperative thin-slice CT was used to classify burr-hole intersection of the underlying MMA groove (no hit, distal-branch hit or main-branch hit) and measure perpendicular burr-hole-to-MMA-groove distance. Co-primary outcomes were radiological recurrence and recurrence requiring intervention. Patient-clustered multivariable logistic regression adjusted for prespecified clinical covariates and treating site.

**Results:** 227 patients (284 operated hemispheres) were included. Radiological recurrence decreased from 34.4% with no branch hit to 22.9% with main-branch intersection, with the gradient confined predominantly to unilateral cSDH. Main-branch intersection was associated with lower adjusted odds of radiological recurrence in unilateral cSDH (adjusted OR 0.30, 95% CI 0.11-0.81; P = .018), with a similar but non-significant association in the overall cohort (adjusted OR 0.53, 95% CI 0.26-1.07; P = .075). Burr-hole-to-MMA-groove distance demonstrated a more consistent association: in the overall cohort, each 5-mm increase independently increased the odds of radiological recurrence (adjusted OR 1.38, 95% CI 1.04-1.82; P = .025). In unilateral cSDH, each 5-mm increase was independently associated with both radiological recurrence (adjusted OR 1.45, 95% CI 1.03-2.04; P = .034) and recurrence requiring intervention (adjusted OR 1.52, 95% CI 1.05-2.20; P = .027).

**Conclusion:** Main-branch intersection of the middle meningeal artery during routine burr-hole surgery is associated with lower recurrence of unilateral cSDH, while the accompanying burr-hole-to-MMA-groove distance gradient provides biologically plausible support for a dose-response relationship. Together, these findings provide mechanistic rationale for prospective evaluation of intentional neuronavigation-guided MMA targeting (BURR-MMA; NCT07549893).

## Introduction

Chronic subdural haematoma (cSDH) is among the most common neurosurgical conditions in older adults, with annual incidence in patients over 65 approaching approximately 50 per 100,000 and rising in parallel with demographic aging and the expanding use of antithrombotic agents^1,2,3^. Burr-hole drainage remains the standard surgical treatment. The fundamental surgical technique, and particularly the anatomical basis for burr-hole placement, has remained largely unchanged since its modern inception. Nevertheless, recurrence remains an important limitation of surgical drainage, occurring in 10-25% of patients across contemporary single- and multicentre series and sometimes necessitating reoperation^4,5^.

The pathophysiology of recurrence is increasingly understood as a vascular phenomenon. The dural neomembrane that develops around chronic subdural collections is densely vascularised, and chronic neovascularisation arising from terminal branches of the middle meningeal artery (MMA) sustains a low-grade rebleed cycle that re-establishes the haematoma following surgical evacuation^6,7^. This mechanistic insight has driven the rapid emergence of endovascular MMA embolisation as an adjunctive treatment. Multiple prospective trials and recent meta-analyses report 50–70% relative reductions in recurrence when embolisation is performed adjunctively or as a stand-alone intervention^8-12^.

A logical extension of this procedure is that intentional intraoperative interruption of the MMA through deliberate burr-hole placement over its calvarial course might confer comparable benefit at the time of standard drainage. Unlike MMA embolisation, optimisation of burr-hole position could potentially target the same vascular pathway without an additional procedure, arterial access, ionising radiation, contrast administration, or specialist endovascular infrastructure. However, whether the anatomical relationship between routinely placed burr holes and the calvarial course of the MMA influences recurrence has not previously been investigated. Standard burr holes are occasionally observed to overlie MMA grooves on the inner table of the skull, but the frequency of this occurrence, its anatomical specificity, and its relationship to recurrence remain unknown.

The distinction between main- and distal-branch intersection is mechanistically important. After entering the cranial vault through the foramen spinosum, the MMA typically divides at or below the pterion into anterior (frontal) and posterior (parietal) main divisions, which subsequently give rise to multiple distal convexity branches supplying the vascularised outer neomembrane^6^. If the therapeutic benefit of MMA-directed treatment derives principally from reducing proximal arterial inflow to this pathological vascular network, burr-hole intersection of the main divisions would be expected to confer the greatest benefit, with progressively smaller effects as intersection occurs more distally.

We therefore performed a multicentre retrospective study to determine whether burr-hole intersection of the MMA is associated with recurrence after cSDH evacuation, and whether any association demonstrates anatomical specificity and a graded distance-response relationship consistent with a biological mechanism.

## Methods

### Study design and ethics

This was a multicentre retrospective cohort study comprising two tertiary central London neurosciences centres: the National Hospital for Neurology and Neurosurgery (NHNN) and the Royal London Hospital (RLH). The study was approved by our local institutional research and development office (Study ID:20241023, R&D Ref ID: 179421 - NHNN), by the clinical audit department (47-202627-SE - RLH) and conducted in accordance with the STROBE guidelines. Given the retrospective observational design and use of routinely collected clinical data, individual informed consent was not required.

### Population

Consecutive adult patients (≥18 years) undergoing burr-hole drainage of cSDH between 1st Jan 2019 and 31st December 2025 at NHNN and RLH were identified from the operative database via the electronic healthcare record (EHR). Patients were eligible if (i) pre- and postoperative CT imaging was available for review and (ii) burr-hole positions could be unambiguously identified relative to MMA groove anatomy. Cases converted to craniotomy were excluded. All operative, imaging, and outcome variables were harmonised to a single common data dictionary, with branch-hit codes (0/1/2), perpendicular distance (millimetres), recurrence definitions, and covariate coding aligned across centres. A binary site indicator (NHNN/RLH) was created for pooled analyses. Sample size was determined pragmatically based on the number of patients available since the inception of the EHR.

### Imaging assessment and exposure classification

Postoperative CT scans (median slice thickness 1.0 mm, range 0.5–1.5 mm; ≤1 mm in 78% of NHNN and 92% of RLH scans) were reviewed in axial, coronal, and sagittal planes. Each burr hole was assessed individually for its relationship to the underlying MMA groove and graded as 0 (no intersection), 1 (intersection of a distal MMA branch), or 2 (intersection of a main MMA division [anterior/frontal or posterior/parietal]), according to the expected calvarial course of the MMA (Figure 1). At the operated-hemisphere level, a main-branch hit was defined by intersection of at least one burr hole with a main MMA division; a distal-only hit required intersection with one or more distal branches without any main-branch intersection. Nearest-groove distance was defined as the minimum perpendicular distance between any burr hole and the nearest MMA groove on the operated hemisphere. Imaging reviewers were blinded to postoperative recurrence and other clinical outcomes during anatomical assessment.

**Figure 1.**
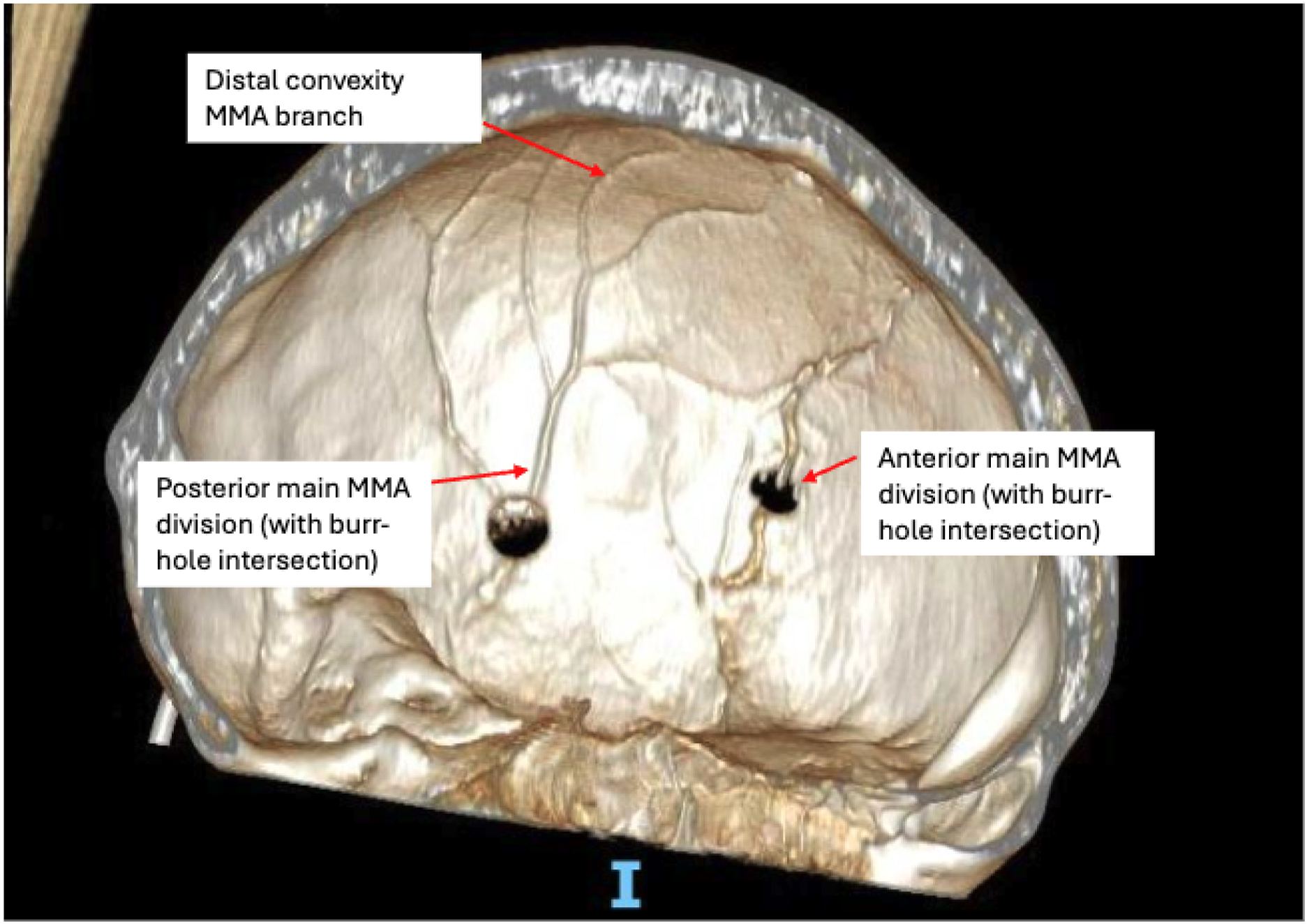
Three-dimensional reconstruction of a bone-window CT scan (1.5-mm slices) demonstrating left-sided burr holes for chronic subdural haematoma evacuation. The calvarial course of the middle meningeal artery (MMA) is shown, with burr-hole intersection of the anterior (frontal) and posterior (parietal) divisions. Distal convexity branches of the MMA are labelled.

The perpendicular distance from the burr-hole edge to the nearest visible MMA groove was recorded in millimetres (Figure 2, 3). Anatomical classification was performed by a single trained reviewer at each centre (TS/JMJ). A random 10% sample underwent independent review by a board-certified consultant neuroradiologist, demonstrating excellent agreement for branch-hit classification (93.5%; Cohen’s κ = 0.89), with no disagreement in main-branch classification.

**Figure 2.**
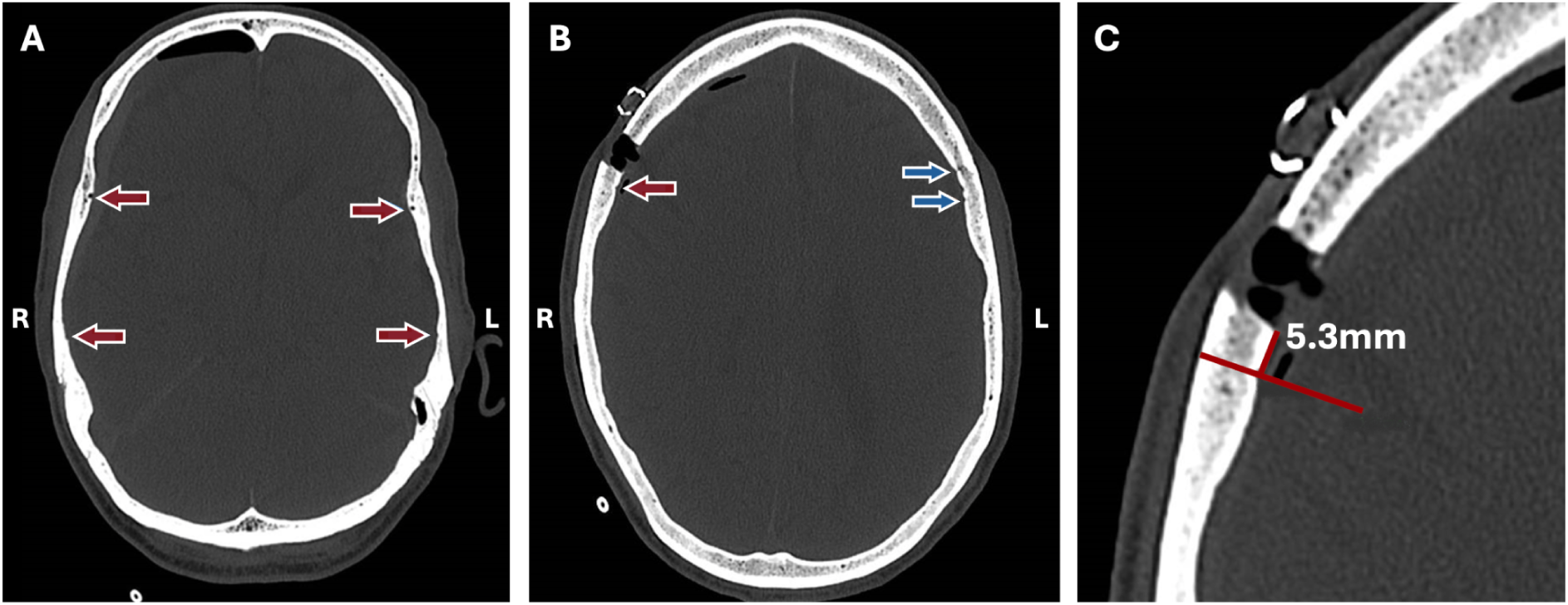
Axial bone window CT images illustrating assessment of burr-hole position relative to the middle meningeal artery (MMA) groove. (A) Representative slice at the level of the pterion demonstrating the four main MMA grooves. (B) Slice at the level of a burr-hole, showing the nearest main groove (red arrow). Distal convexity branches arising from the contralateral anterior division are shown for reference (blue arrows). (C) Measurement of the perpendicular distance from the burr-hole edge to the nearest visible MMA groove (example shown: 5.3 mm).

**Figure 3.**
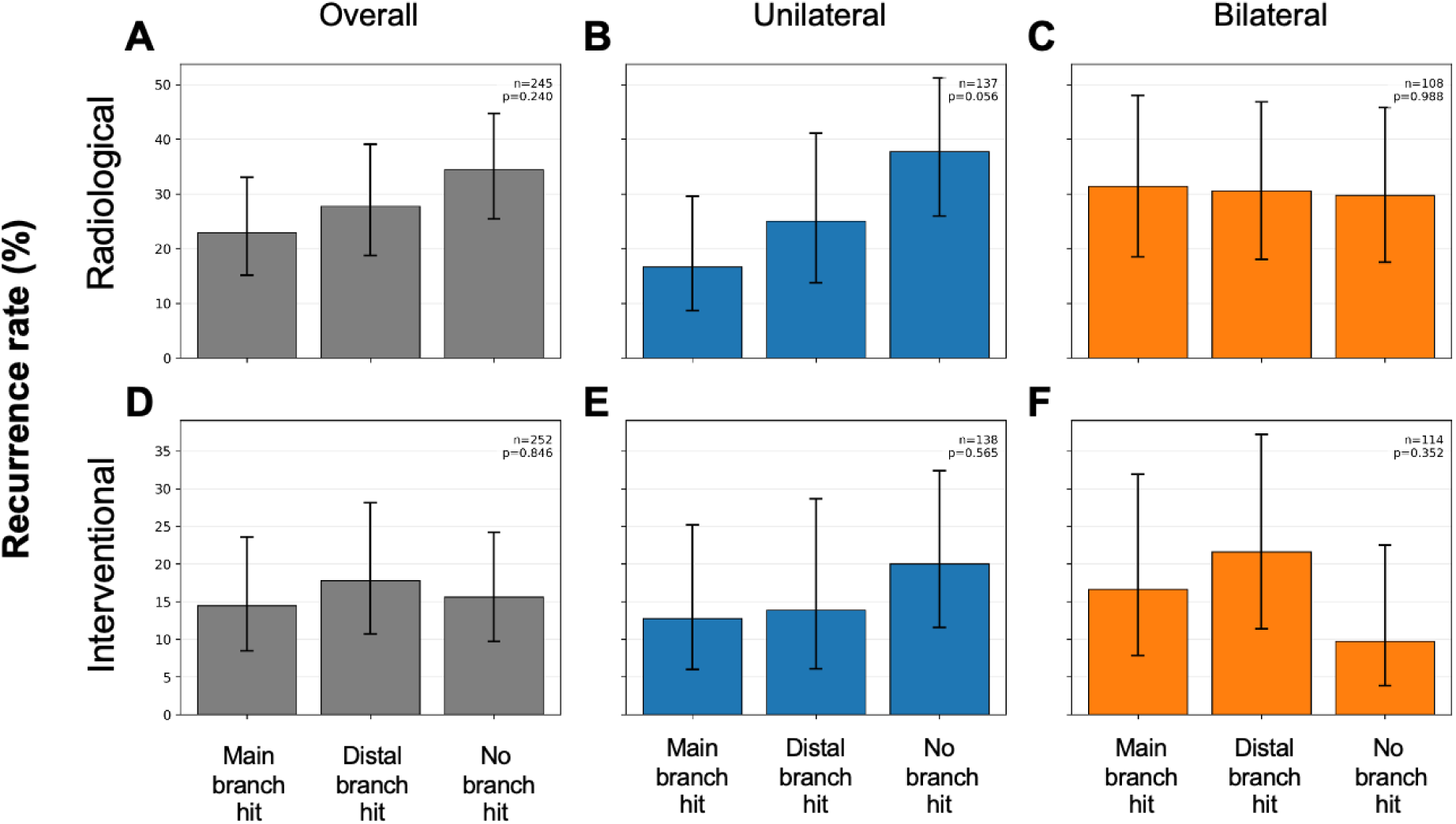
Radiological (A-C) and Interventional (D-F) rate of recurrence compared by branch hit group. Global significance values are reported across the group.

The pre-specified primary exposure was a hemisphere-level binary variable, Main branch hit (score of 2). Pre-specified secondary exposures comprised a three-level categorical variable (Group A = main hit; Group B = distal-only hit; Group C = no hit, with Group C as reference); a count of main-branch hits per hemisphere (0-2); a binary “any branch hit” variable; and the continuous burr-hole-to-MMA-groove distance in millimetres.

### Data and statistical analysis

The co-primary outcomes were (i) radiological recurrence, defined as a new or enlarging ipsilateral subdural collection on follow-up imaging, and (ii) recurrence requiring intervention, defined as re-operation for recurrent ipsilateral cSDH. Secondary outcome was favourable functional outcome at 3-6 months (modified Rankin Scale 0–2).

Baseline characteristics were summarised overall and stratified by main-branch intersection status. Continuous variables are presented as median (interquartile range [IQR]) and categorical variables as frequency (%). Group comparisons were performed using the Mann–Whitney U or Kruskal–Wallis test for continuous variables, and the χ² or Fisher’s exact test for categorical variables, as appropriate. Nearest-groove distance was categorised in 5-mm intervals to provide clinically interpretable spatial groupings, minimise false precision arising from CT-based measurement variability, and allow assessment of a potentially non-linear association with recurrence.

The primary analysis evaluated the association between main-branch intersection and recurrence in the combined cohort of operated hemispheres using multivariable logistic regression. As patients with bilateral cSDH could contribute two operated hemispheres, observations from the same patient were not considered statistically independent. Patient-level cluster-robust standard errors were therefore calculated using a sandwich variance estimator, with patient ID defining the clusters. Models were presented unadjusted and adjusted a priori for age, sex, maximal haematoma thickness, prior anticoagulant or antiplatelet use, bilateral status and treating site. Adjusted odds ratios (ORs) with 95% confidence intervals (CIs) were reported.

Pre-specified sensitivity analyses included: (i) exclusion of bilateral cSDH cases and (ii) Cox proportional hazards modelling for interventional recurrence specifically, where time-to-event data were available. All analyses were performed using Python (version 3.11.15). Statistical tests were two-sided, with P < .05 considered statistically significant. Analyses were performed using complete cases for the outcome, exposure, and covariates included in each model. Consequently, denominators varied slightly between outcomes according to data availability. Models that were directly compared, including nested models and analyses assessing attenuation after inclusion of recurrence, were fitted using an identical common complete-case cohort. The numbers of operated hemispheres, unique patients, and outcome events included in each model are reported. Where model fit was compared, comparisons were restricted to models fitted to the same outcome and observations.

### Data Availability

The data underlying this study contain potentially identifiable clinical information and are not publicly available. Deidentified data may be made available by the corresponding author upon reasonable request, subject to institutional approval and applicable data-governance requirements.

## Results

### Patients and centres

227 patients were operated on across both centres (Table 1) representing 284 operated hemispheres. Of these, 72 patients (32%) had bilateral SDH, although not all underwent bilateral operations. Subdural collection thickness was lower at RLH than at NHNN, and there were fewer female patients at RLH (Table 1). No significant differences were found in pre-operative GCS or existing co-morbidities such as hypertension and diabetes, nor in the use of blood thinners.

**Table 1.** Values are presented as *n* (%), *n/N* (%), or median [interquartile range]. SMD denotes standardised mean difference. Bold indicates *p*<0.05.

| Category | Variable | Overall | NHNN | RLH | p | Test | SMD |
| --- | --- | --- | --- | --- | --- | --- | --- |
| **Demographics** | Number of patients | 227 | 190 | 37 | — | — | — |
| | Female sex | 77 (33.9%) | 69 (36.3%) | 8 (21.6%) | 0.084 | $\chi^2$ | 0.328 |
|  | Age, years | 74.0<br>[64.5–82.0] | 74.0<br>[65.0–81.0] | 73.0<br>[60.0–88.0] | 0.542 | MWU | −0.045 |
|  | Preoperative GCS | 14.0<br>[14.0–15.0] | 14.0<br>[14.0–15.0] | 14.0<br>[14.0–15.0] | 0.300 | MWU | −0.282 |
| **Subdural characteristics** | Maximum haematoma thickness, mm | 22.8<br>[17.1–27.0] | 23.0<br>[18.0–28.0] | 17.4<br>[15.0–25.0] | **0.002** | MWU | 0.522 |
|  | Midline shift, mm | 8.0<br>[4.8–11.9] | 8.5<br>[5.0–12.0] | 7.0<br>[3.6–10.0] | 0.118 | MWU | 0.311 |
| **Comorbidities** | Hypertension | 122/227<br>(53.7%) | 105/190<br>(55.3%) | 17/37<br>(45.9%) | 0.298 | $\chi^2$ | 0.187 |
| | Diabetes mellitus | 62/227<br>(27.3%) | 53/190<br>(27.9%) | 9/37 (24.3%) | 0.656 | $\chi^2$ | 0.081 |
|  | Chronic lung disease | 9/227<br>(4.0%) | 7/190<br>(3.7%) | 2/37 (5.4%) | 0.643 | FE | −0.083 |
| | Kidney disease | 34/227<br>(15.0%) | 29/190<br>(15.3%) | 5/37 (13.5%) | 0.785 | $\chi^2$ | 0.050 |
|  | Atrial fibrillation | 27/227<br>(11.9%) | 24/190<br>(12.6%) | 3/37 (8.1%) | 0.584 | FE | 0.149 |
| | Prior anticoagulant or antiplatelet use | 77/227<br>(33.9%) | 61/190<br>(32.1%) | 16/37<br>(43.2%) | 0.191 | $\chi^2$ | −0.231 |
| | Bilateral cSDH | 72/227<br>(31.7%) | 61/190<br>(32.1%) | 11/37<br>(29.7%) | 0.776 | $\chi^2$ | 0.051 |
| **Surgical characteristics** | Nearest groove distance, mm | 0.0<br>[0.0–3.7] | 0.0<br>[0.0–3.3] | 0.0 [0.0–5.0] | 0.907 | MWU | −0.246 |
|  | Burr holes per hemisphere | 2.0<br>[2.0–2.0] | 2.0<br>[2.0–2.0] | 2.0 [2.0–2.0] | 0.581 | MWU | –0.087 |
|  | Drain inserted, per hemisphere | 266/281<br>(94.7%) | 218/233<br>(93.6%) | 48/48<br>(100.0%) | 0.082 | FE | –0.371 |
| | Branch-hit group: none / distal / main, per hemisphere | 108 / 80 / 95 | 92 / 61 / 82 | 16 / 19 / 13 | 0.158 | $\chi^2$ | — |

### Radiological and interventional recurrence by group and nearest groove distance

Median follow-up was 84 days (IQR 48-172 days). Radiological recurrence decreased from 34.4% with no branch hit to 27.8% with a distal convexity branch-only hit and 22.9% with a main-branch hit, although the overall difference was not significant (χ²(2)=2.86, P = .240) [Figure 3A]. When stratified by laterality, this gradient was confined to unilateral cSDH, in which recurrence was 37.7% with no hit, 25.0% with a distal-branch hit and 16.7% with a main-branch hit (global P = .056) [Figure 3B]; the main-branch versus no-hit comparison was significant (OR 0.33, 95% CI 0.13–0.85; P = .026). In bilateral cSDH, radiological recurrence was similar across main-branch, distal-branch and no-hit groups (31.4%, 30.6% and 29.7%, respectively; P = .988) [Figure 3C]. Intervention recurrence did not differ between branch-hit groups overall (15.6%, 17.8% and 14.5% for no hit, distal-only hit and main-branch hit, respectively; χ²(2)=0.33, P = .846) [Figure 3D], nor within unilateral (20.0%, 13.9% and 12.8%; P = .565) [Figure 3E] or bilateral cohorts (9.8%, 21.6% and 16.7%; P = .352) [Figure 3F].

Nearest-groove distance was not significantly associated with radiological recurrence in the overall cohort (unadjusted OR 1.28 per 5-mm increase, 95% CI 0.97–1.69; P = .076) [Figure 4A]. In unilateral cSDH, increasing nearest-groove distance was associated with radiological recurrence (unadjusted OR 1.42 per 5 mm, 95% CI 1.01–1.98; P = .042) [Figure 4B], whereas no corresponding association was observed in bilateral cSDH (unadjusted OR 1.03 per 5 mm; P = .906) [Figure 4C]. For recurrence requiring intervention, nearest-groove distance was not significantly associated with recurrence in the overall cohort (unadjusted OR 1.31 per 5-mm increase, 95% CI 0.98–1.77; P = .072) [Figure 4D]. However, increasing distance was associated with recurrence requiring intervention in unilateral cSDH (unadjusted OR 1.50 per 5 mm, 95% CI 1.05–2.12; P = .024) [Figure 4E], while no corresponding association was observed in bilateral cSDH (unadjusted OR 0.82 per 5 mm; P = .626) [Figure 4F].

**Figure 4.**
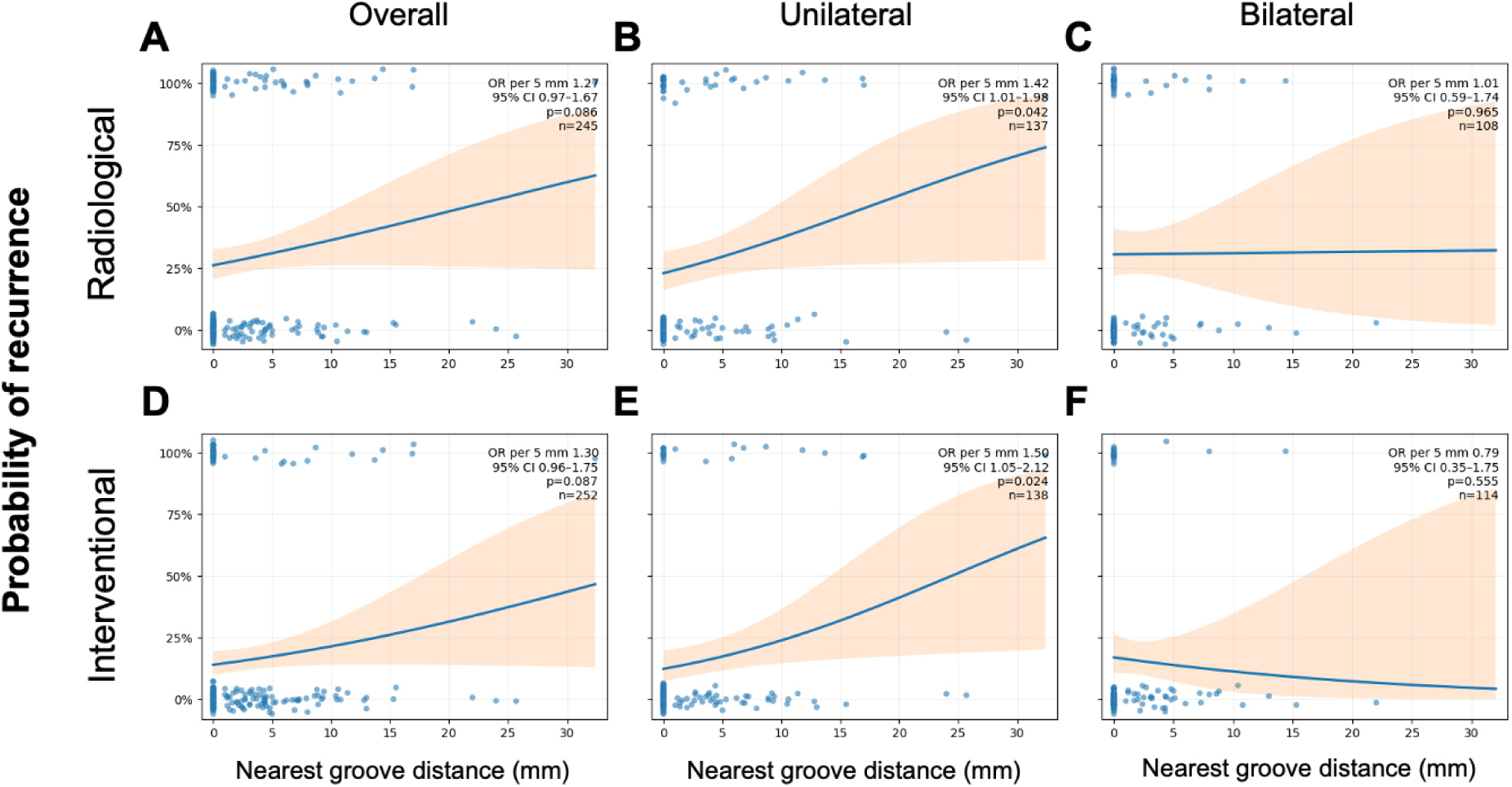
Association between nearest-groove distance and recurrence according to cSDH laterality. Jittered binary outcome plots with fitted univariable logistic-regression curves show the association between nearest middle meningeal artery groove distance and radiological recurrence (A–C) recurrence requiring intervention (D–F), for the overall cohort, unilateral cSDH, and bilateral cSDH, respectively. Odds ratios are reported per 5-mm increase in nearest-groove distance. Points represent individual operated hemispheres, and solid lines represent model-predicted recurrence probabilities with orange shaded 95% confidence intervals.

### Patient-clustered logistic regression models by group and nearest groove distance

Given the potential influence of clinical and procedural confounders, and the non-independence of observations from patients with bilateral cSDH, separate patient-clustered logistic regression models were fitted for each exposure as previously described (Table 2). Only complete cases were included in the multivariate analyses. In this way, 242 and 248 hemispheres were retained for radiological and interventional recurrence respectively. In the patient-clustered multivariable model for radiological recurrence, neither distal-only branch hit (adjusted OR 0.69, 95% CI 0.33–1.44; P = .320) nor main-branch hit (adjusted OR 0.53, 95% CI 0.26–1.07; P = .075) was significantly associated with recurrence compared with no branch hit. None of the prespecified covariates were independently associated with radiological recurrence, although there were non-significant trends towards lower recurrence with main-branch hit and treatment at RLH.

**Table 2.**
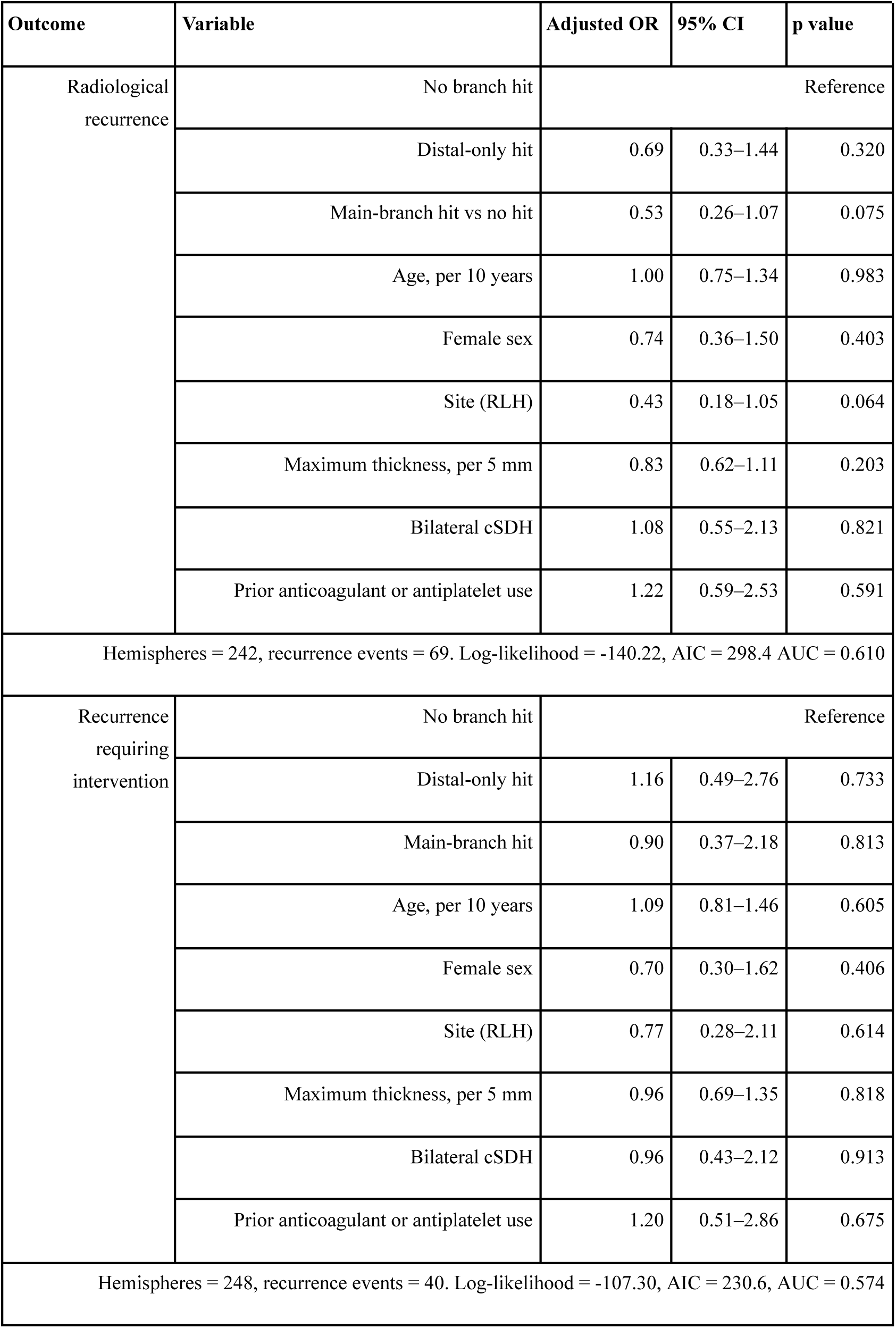
Patient-clustered multivariable logistic regression of branch-hit group and radiological and interventional recurrence. The intercept was included in the model but omitted from the table for brevity. AIC = Akaike Information Criterion; AUC = area under curve; cSDH = chronic subdural haematoma; RLH = Royal London Hospital.

| Outcome | Variable | Adjusted OR | 95% CI | p value |
| --- | --- | --- | --- | --- |
| Radiological recurrence | No branch hit | Reference |  |  |
|  | Distal-only hit | 0.69 | 0.33–1.44 | 0.320 |
|  | Main-branch hit vs no hit | 0.53 | 0.26–1.07 | 0.075 |
|  | Age, per 10 years | 1.00 | 0.75–1.34 | 0.983 |
|  | Female sex | 0.74 | 0.36–1.50 | 0.403 |
|  | Site (RLH) | 0.43 | 0.18–1.05 | 0.064 |
|  | Maximum thickness, per 5 mm | 0.83 | 0.62–1.11 | 0.203 |
|  | Bilateral cSDH | 1.08 | 0.55–2.13 | 0.821 |
|  | Prior anticoagulant or antiplatelet use | 1.22 | 0.59–2.53 | 0.591 |
| Hemispheres = 242, recurrence events = 69. Log-likelihood = -140.22, AIC = 298.4 AUC = 0.610 |  |  |  |  |
| Recurrence requiring intervention | No branch hit | Reference |  |  |
|  | Distal-only hit | 1.16 | 0.49–2.76 | 0.733 |
|  | Main-branch hit | 0.90 | 0.37–2.18 | 0.813 |
|  | Age, per 10 years | 1.09 | 0.81–1.46 | 0.605 |
|  | Female sex | 0.70 | 0.30–1.62 | 0.406 |
|  | Site (RLH) | 0.77 | 0.28–2.11 | 0.614 |
|  | Maximum thickness, per 5 mm | 0.96 | 0.69–1.35 | 0.818 |
|  | Bilateral cSDH | 0.96 | 0.43–2.12 | 0.913 |
|  | Prior anticoagulant or antiplatelet use | 1.20 | 0.51–2.86 | 0.675 |
| Hemispheres = 248, recurrence events = 40. Log-likelihood = -107.30, AIC = 230.6, AUC = 0.574 |  |  |  |  |

**Table 3.** Patient-clustered multivariable logistic regression of nearest groove distance and radiological and interventional recurrence. The intercept was included in the model but omitted from the table for brevity. AIC = Akaike Information Criterion; AUC = area under curve; cSDH = chronic subdural haematoma; RLH = Royal London Hospital.

| Outcome | Variable | Adjusted OR | 95% CI | p value |
| --- | --- | --- | --- | --- |
| Radiological recurrence | Nearest-groove distance, per 5mm | 1.38 | 1.04–1.82 | **0.025** |
|  | Female sex | 0.73 | 0.36–1.50 | 0.392 |
|  | Site (RLH) | 0.39 | 0.16–0.97 | **0.043** |
|  | Maximum thickness, per 5 mm | 0.83 | 0.62–1.12 | 0.218 |
|  | Bilateral cSDH | 1.16 | 0.59–2.27 | 0.669 |
|  | Prior anticoagulant or antiplatelet use | 1.28 | 0.62–2.66 | 0.506 |
|  | Age, per 10 years | 0.99 | 0.74–1.32 | 0.948 |
| Hemispheres = 242, recurrence events = 69. Log-likelihood = -139.57, AIC = 295.14, AUC = 0.607 |  |  |  |  |
| Recurrence requiring intervention | Nearest-groove distance, per 5mm | 1.33 | 0.97–1.82 | 0.074 |
|  | Female sex | 0.69 | 0.30–1.60 | 0.390 |
|  | Site (RLH) | 0.71 | 0.25–1.96 | 0.504 |
|  | Maximum thickness, per 5 mm | 0.94 | 0.67–1.32 | 0.714 |
|  | Bilateral cSDH | 1.04 | 0.47–2.30 | 0.930 |
|  | Prior anticoagulant or antiplatelet use | 1.28 | 0.53–3.06 | 0.582 |
|  | Age, per 10 years | 1.08 | 0.80–1.46 | 0.605 |
| Hemispheres = 248, recurrence events = 40. Log-likelihood = -107.30, AIC = 232.6, AUC = 0.574 |  |  |  |  |

### Bilateral cSDH-excluded sensitivity analyses

A sensitivity analysis was performed after excluding all patients with bilateral cSDH (Supplemental Tables 1 and 2). As each remaining patient contributed only one operated-hemisphere observation, conventional multivariable binary logistic regression was used without patient-level clustering. Here, main-branch hit was associated with lower odds of radiological recurrence (adjusted OR 0.30, 95% CI 0.11–0.81; P = .018), whereas the association for distal-only hit was not significant (adjusted OR 0.52, 95% CI 0.19–1.39; P = .191). Increasing nearest-groove distance was independently associated with both outcomes. Each 5-mm increase was associated with a 45% increase in the adjusted odds of radiological recurrence (adjusted OR 1.45, 95% CI 1.03–2.04; P = .034) and a 52% increase in the odds of recurrence requiring intervention (adjusted OR 1.52, 95% CI 1.05–2.20; P = .027). Notably for all 4 models, discrimination as measured by AUC substantially improved as compared to the bilateral-included models (Supplementary Tables 1 and 2).

### Time-to-event sensitivity analyses

The median time to recurrence intervention was 17 days (IQR: 9-38). Patient-clustered Cox proportional-hazard models were fit using time from surgery to interventional recurrence, with adjustment for covariates. Hemispheres without recurrence were censored at their last recorded follow-up; up to 6 months. Again, 248 hemispheres were available for this analysis. Branch-hit group was not associated with recurrence requiring intervention. Compared with no branch hit, distal-only branch hit was associated with an adjusted hazard ratio of 1.09 (95% CI 0.51–2.34; P = .823), while main-branch hit was associated with an adjusted hazard ratio of 0.88 (95% CI 0.39–2.03; P = .773). The global patient-clustered Wald test was non-significant (χ²(2)=0.27; P = .876), as was the complementary partial likelihood-ratio test (χ²(2)=0.27; P = .874). Kaplan–Meier curves were similar across branch-hit groups, with no evidence of separation on unadjusted log-rank testing (χ²(2)=0.26; P = .879) [Figure 5A].

**Figure 5.**
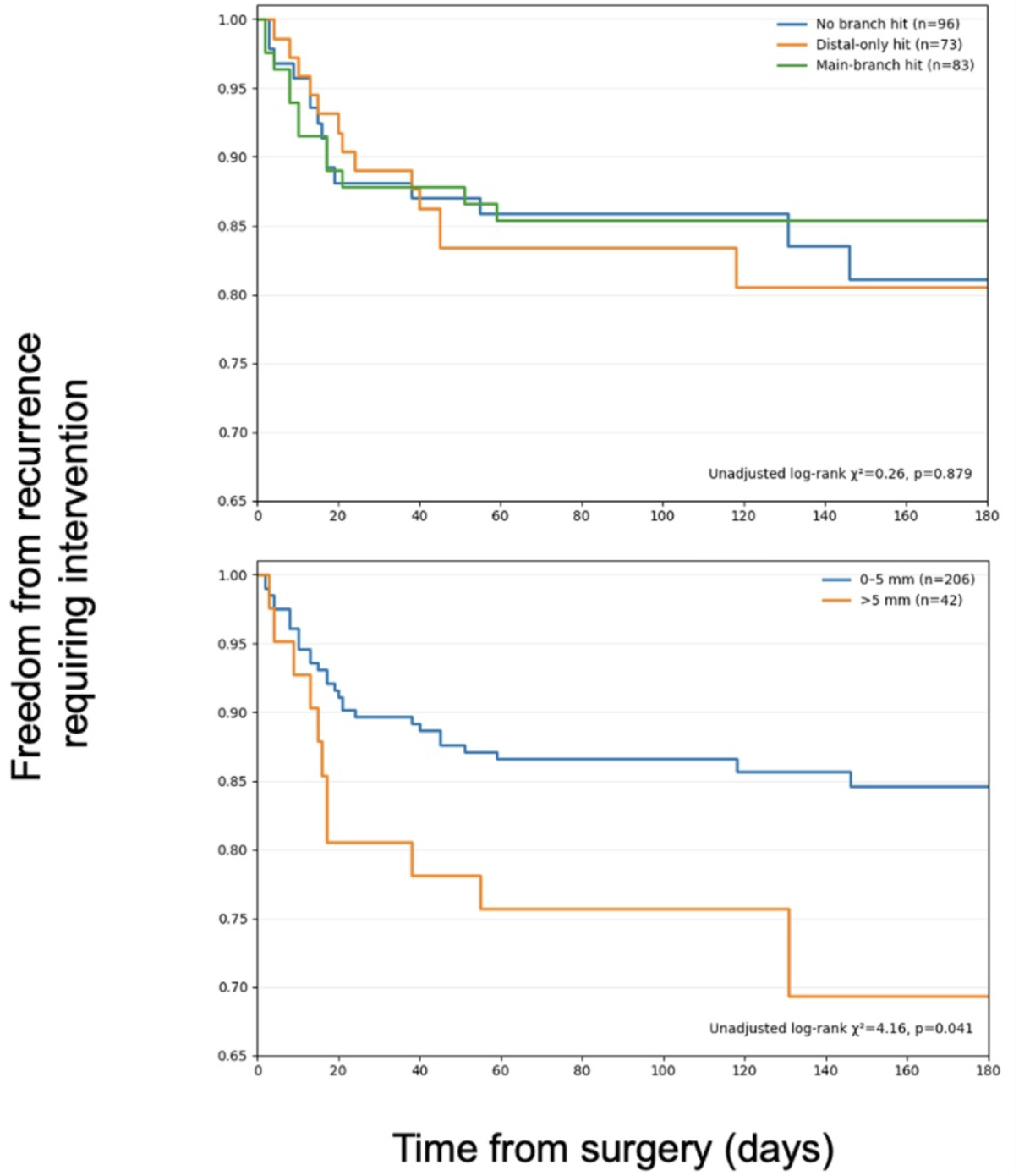
Kaplan–Meier estimates of freedom from recurrence requiring intervention within 180 days. (5A) Freedom from recurrence requiring intervention according to branch-hit group, comparing no branch hit, distal-only branch hit, and main-branch hit. (5B) Freedom from recurrence requiring intervention according to dichotomised nearest-groove distance.

Nearest-groove distance was dichotomised at 5 mm to distinguish burr holes placed within close proximity to the groove from those positioned further away. Compared with distances of 0–5 mm, distances >5 mm were associated with a higher hazard of recurrence requiring intervention at or before 180 days (adjusted HR 2.22, 95% CI 1.05–4.70; P = .037) [Figure 5B]. Alternative nearest-groove distance categorisations were explored, and model fit was compared using the patient-clustered global Wald test, partial likelihood-ratio test, AIC, and Harrell C-index; the 0–5 mm versus >5 mm specification provided the most parsimonious and statistically coherent fit.

### Influence of burr-hole nearest groove distance on mRS

Each 5-mm increase in nearest-groove distance was associated with higher odds of unfavourable functional outcome (mRS 3-6) on unadjusted analysis (OR 1.40, 95% CI 1.06–1.84; P = .016), with a similar but non-significant adjusted estimate (adjusted OR 1.45, 95% CI 0.99–2.12; P = .060). Further adjustment for recurrence attenuated this association (adjusted OR 1.37, 95% CI 0.92–2.05; P = .126), while recurrence itself was independently associated with unfavourable mRS (adjusted OR 3.06, 95% CI 1.16–8.03; P = .023). These findings may be consistent with recurrence partially accounting for the association between greater nearest-groove distance and worse functional outcome, although the evidence remains exploratory.

## Discussion

This multicentre study demonstrates that the anatomical relationship between burr-hole position and the middle meningeal artery (MMA) is associated with postoperative recurrence following burr-hole drainage for chronic subdural haematoma. Three observations underpin this conclusion: first, the protective association was concentrated in burr holes intersecting the main MMA divisions, suggesting anatomical specificity; second, recurrence increased progressively with increasing burr-hole-to-MMA-groove distance, consistent with a biological gradient; and third, these findings were reproduced across two independent centres and were most evident in unilateral disease. Together, these observations support a biologically plausible relationship between surgical anatomy and recurrence.

### Biological plausibility and mechanistic interpretation

Anatomical specificity is an important feature of these findings. The protective association was concentrated in burr holes intersecting the main MMA divisions, whereas distal-branch intersection alone was not associated with lower recurrence. This pattern accords with the known arterial anatomy of the MMA. Following entry through the foramen spinosum, the anterior and posterior main divisions provide the principal inflow to the distal convexity branches supplying the vascularised outer neomembrane. Disruption of these proximal divisions would therefore be expected to have a greater effect on downstream neomembrane perfusion than disruption of individual distal branches^13,14^.

Recurrence is now understood as an active vascular and inflammatory process sustained by fragile neovascular channels within the outer neomembrane, supplied predominantly by distal convexity branches arising from the anterior and posterior divisions of the MMA^6,7^. Interruption of this arterial supply, whether by endovascular embolisation, deliberate surgical interruption, or, as these findings suggest, incidental disruption during routine burr-hole surgery, may reduce neomembrane perfusion, attenuate the inflammatory cycle, and lower the risk of recurrent haematoma formation. Burr holes placed directly over or immediately adjacent to the calvarial groove of a main MMA division may partially disrupt the vessel or its accompanying dural branches during drilling, dural opening, bipolar coagulation of the dural edge, or drain placement.

Within this biological framework, burr-hole-to-MMA-groove distance emerged as the most informative anatomical metric. Each 5-mm increase in distance was independently associated with higher odds of radiological recurrence, with a similar direction of effect for recurrence requiring intervention. In unilateral cSDH, increasing distance independently predicted both radiological and intervention-requiring recurrence, while burr holes located more than 5 mm from the nearest groove were associated with more than twice the hazard of recurrence requiring intervention. Greater groove distance was also associated with worse functional outcome, although this relationship attenuated after adjustment and after accounting for recurrence, suggesting that recurrence may partially mediate any effect on disability.

The superiority of the continuous distance metric over binary branch intersection is itself biologically plausible. The calvarial groove is a surrogate for the underlying artery, and the likelihood or extent of arterial disruption during drilling and dural opening is unlikely to be an all-or-none phenomenon. Rather, anatomical proximity probably represents a continuum in which progressively closer burr-hole placement may increase the likelihood or extent of arterial or dural vascular disruption. This interpretation is consistent with contemporary models of cSDH pathophysiology and with the biological gradient observed in the present study. Together with the established efficacy of MMA embolisation^8-12^ and recent reports of deliberate intraoperative MMA interruption^15,16^, these findings support a coherent mechanistic hypothesis rather than an isolated statistical association. Nevertheless, causality cannot be inferred from this observational study, which exploits naturally occurring anatomical variation during otherwise standardised burr-hole surgery.

### Position within the current MMA-directed evidence base

Randomised trials of MMA embolisation, including MAGIC-MT, EMBOLISE and STEM, together enrolling more than 1,400 patients, have established MMA-directed therapy as an effective strategy for reducing cSDH recurrence^8-12^. A recent meta-analysis reported an approximately 50% relative reduction in recurrence or progression and a significant reduction in 90-day rescue surgery, with directionally consistent effects in surgically treated patients^8^. These findings establish the MMA as a therapeutically relevant target in cSDH.

The present study does not compare with or seek to replace MMA embolisation, but addresses a complementary mechanistic question: whether the anatomical relationship between routinely placed burr holes and the MMA influences recurrence. Because no deliberate MMA targeting was undertaken, routine surgery provides a naturally occurring experiment in which variation in burr-hole position may alter the likelihood of arterial or dural vascular disruption. Our findings suggest that this previously overlooked component of operative technique may influence recurrence and warrants prospective evaluation.

Recent studies of intentional surgical MMA interruption support this concept. Intentional surgical MMA interruption provides further support for this hypothesis. Sun et al. randomised 72 patients to MMA-targeted burr-hole placement with intraoperative coagulation versus conventional drainage, reporting no recurrences in the intervention arm and reductions in inflammatory and angiogenic mediators^15^. Dowaki et al. subsequently reported lower recurrence with three-dimensional CT-guided burr-hole MMA coagulation (3.2% vs 15.8%)^16^. The present study complements these reports in three important ways. First, it demonstrates anatomical specificity, with the strongest association observed when the main MMA divisions are intersected. Second, it provides the first quantitative burr-hole-to-MMA-groove distance-response relationship, suggesting that the effect is graded rather than binary. Third, because no deliberate MMA targeting was performed, the observed association arises entirely from naturally occurring anatomical variation during standard surgery. This likely represents a conservative estimate of the potential effect achievable through intentional neuronavigation-guided MMA targeting. Although MMA embolisation aims to achieve distal penetration of embolic material into the pathological neomembrane vasculature, this is accomplished through catheterisation of the anterior or posterior MMA divisions supplying the convexity dura^14,17-19^. Our findings suggest that interruption of these proximal divisions may itself influence recurrence, consistent with the concept that reducing inflow to the distal vascular network, rather than directly treating individual neovessels, is the critical therapeutic principle. The observation that main-division intersection was associated with lower recurrence, whereas distal-only intersection was not, further supports this anatomical hierarchy.

### The unilateral-bilateral divergence

The protective association was largely confined to unilateral cSDH and was attenuated in bilateral disease. Several biological explanations are plausible. Bilateral cSDH may represent a clinically distinct phenotype, with previous surgical series identifying bilateral disease as a predictor of recurrence. Whether this reflects differences in neomembrane biology, vascular supply, brain re-expansion, or other patient-level factors remains uncertain. One possibility is that a more distributed pathological process could reduce the relative effect of focal interaction with a single MMA territory, although this remains speculative^20-21^.

Equally, methodological factors may have contributed. Patients with bilateral cSDH contribute two operated hemispheres with correlated outcomes, and although patient-level cluster-robust variance estimation accounts for statistical dependence, it does not fully address the complexities of attributing recurrence to hemisphere-level exposures. Furthermore, the relatively small number of bilateral recurrence events limited precision. These findings should not be interpreted as evidence that bilateral cSDH is unresponsive to MMA-directed therapy; bilateral MMA embolisation has been reported with favourable outcomes, although dedicated evidence in this subgroup remains limited^22^. Rather, they suggest that bilateral disease may warrant separate biological and statistical consideration. Future prospective studies should pre-specify unilateral and bilateral cohorts rather than combining them within a single analytical framework.

### Clinical implications

The median burr-hole-to-groove distance was zero, and most burr holes lay within a few millimetres of a groove; the shift required to convert a "near-miss" into a main-branch intersection is small and would fall entirely within the standard operative field. Modest optimisation of burr-hole placement, informed by the pre-operative CT already performed as routine care, is biologically rational and operationally deliverable in any NHS neurosurgical unit without new equipment. Neuronavigation provides a natural mechanism for this optimisation and records the achieved position for prospective fidelity monitoring. These considerations do not license immediate practice change; they provide a defensible mechanistic and quantitative rationale for the ongoing BURR-MMA prospective trial (NCT07549893).

### Strengths and Limitations

This multicentre study used thin-slice CT to characterise burr-hole position using both branch classification and continuous burr-hole-to-MMA-groove distance, allowing assessment of anatomical specificity and biological gradient. Exposure classification was harmonised across centres and underwent independent quality-control review, including consultant neuroradiology adjudication. Patient-clustered models accounted for within-patient correlation in bilateral disease, and both radiological and intervention-requiring recurrence were evaluated.

Several limitations should be acknowledged. Burr-hole position was not randomly assigned, leaving the possibility of residual confounding. Radiological follow-up was not protocolised and was often symptom-triggered, potentially underestimating asymptomatic recurrence. The second-centre cohort was modest, limiting adjusted estimation of the binary exposure, although continuous-distance analyses remained interpretable. Exposure classification was performed by a single trained reviewer at each centre without formal cross-institutional inter-rater assessment. Hemisphere-level analysis remains imperfect in bilateral disease despite patient-clustered modelling, and intervention-recurrence events were relatively few. Finally, no intentional MMA interruption was performed; these findings reflect naturally occurring anatomical variation rather than a therapeutic intervention and therefore cannot establish causality. Prospective evaluation with intentional targeting and protocolised follow-up is required.

## Conclusions

In this multicentre retrospective cohort, burr-hole intersection of a middle meningeal artery main branch was associated with a substantial reduction in radiological recurrence of chronic subdural haematoma, with external validation in an independent cohort and statistically significant strengthening on pooled multicentre analysis. The protective effect was anatomically specific to main-branch intersection and was independently corroborated by a continuous-distance gradient. Because the majority of patients were already within 5 mm of an MMA groove, only modest refinement of standard burr-hole placement is required to translate these findings into routine practice. These data provide a mechanistically coherent, externally validated, hypothesis-strengthening rationale for a prospective trial of intentional MMA main-branch targeting (BURR-MMA, <u>clinicaltrials.gov</u> NCT07549893).

## Supporting information

Supplementary Results

## References

1. Yang W, Huang J. Chronic subdural hematoma: epidemiology and natural history. Neurosurg Clin N Am. 2017;28(2):205–210.

2. Dziho A, et al. Global prevalence and incidence of chronic subdural hematoma: a systematic review. Brain Spine. 2025;5:105893. doi:10.1016/j.bas.2025.105893.

3. Stubbs DJ, Vivian ME, Davies BM, Ercole A, Burnstein R, Joannides AJ. Incidence of chronic subdural haematoma: a single-centre exploration of the effects of an ageing population with a review of the literature. Acta Neurochir (Wien). 2021;163(9):2629–2637. doi:10.1007/s00701-021-04879-z.

4. Almenawer SA, Farrokhyar F, Hong C, et al. Chronic subdural hematoma management: a systematic review and meta-analysis of 34,829 patients. Ann Surg. 2014;259(3):449–457. doi:10.1097/SLA.0000000000000255.

5. Santarius T, Kirkpatrick PJ, Ganesan D, et al. Use of drains versus no drains after burr-hole evacuation of chronic subdural haematoma: a randomised controlled trial. Lancet. 2009;374(9695):1067–1073. Doi:10.

6. 1016/S0140-6736(09)61115-6.

7. Edlmann E, Giorgi-Coll S, Whitfield PC, Carpenter KLH, Hutchinson PJ. Pathophysiology of chronic subdural haematoma: inflammation, angiogenesis and implications for pharmacotherapy. J Neuroinflammation. 2017;14:108. doi:10.1186/s12974-017-0881-y.

8. Tanaka T, Kaimori M. Histological study of vascular structure between the dura mater and the outer membrane in chronic subdural hematoma in an adult. No Shinkei Geka. 1999;27(5):431-436.

9. Gillespie CS, Veremu M, Cook WH, et al. Middle meningeal artery embolization for chronic subdural hematoma: meta-analysis of three randomized controlled trials and review of ongoing trials. Acta Neurochir (Wien). 2025;167(1):166. doi:10.1007/s00701-025-06587-4.

10. Davies JM, Knopman J, Mokin M, et al; EMBOLISE Investigators. Adjunctive middle meningeal artery embolization for subdural hematoma. N Engl J Med. 2024;391(20):1890–1900. doi:10.1056/NEJMoa2313472.

11. Fiorella D, Monteith SJ, Hanel R, et al; STEM Investigators. Embolization of the middle meningeal artery for chronic subdural hematoma. N Engl J Med. 2025;392(9):855–864. doi:10.1056/NEJMoa2409845.

12. Liu J, Ni W, Zuo Q, et al. Middle meningeal artery embolization for nonacute subdural hematoma. N Engl J Med. 2024;391(20):1901–1912. doi:10.1056/NEJMoa2401201.

13. Srivatsan A, Mohanty A, Nascimento FA, et al. Middle meningeal artery embolization for chronic subdural hematoma: meta-analysis and systematic review. World Neurosurg. 2019;122:613–619. doi:10.1016/j.wneu.2018.11.167.

14. Mino M, Nishimura S, Hori E, et al. Efficacy of middle meningeal artery embolization in the treatment of refractory chronic subdural hematoma. Surg Neurol Int. 2010;1:78. doi:10.4103/2152-7806.73801.

15. Shapiro M, Walker M, Carroll KT, et al. Neuroanatomy of cranial dural vessels: implications for subdural hematoma embolization. J Neurointerv Surg. 2021;13(5):471–477.

16. Sun T, Shao D, Li J, et al. Therapeutic efficacy of drilling drainage combined with intraoperative middle meningeal artery occlusion in the management of chronic subdural hematoma: a clinical study. Neurosurg Rev. 2024;47(1):293. doi:10.1007/s10143-024-02501-1.

17. Dowaki R, Fukuda S, Taniguchi H, Watanabe Y, Horie N. Three-dimensional computed tomography-guided burr-hole surgery with middle meningeal artery coagulation and severance (B-MACS) for chronic subdural hematoma: a retrospective clinical study. Neurosurg Rev. 2026;49(1):336. doi:10.1007/s10143-026-04265-2.

18. Link TW, Rapoport BI, Paine SM, Kamel H, Knopman J. Middle meningeal artery embolization for chronic subdural hematoma: endovascular technique and radiographic findings. Interv Neuroradiol. 2018;24(4):455–462. doi:10.1177/1591019918769336.

19. Siddiq F, Shakir M, Nguyen TN, et al. Consensus statement on middle meningeal artery embolization in chronic subdural hematoma treatment: a guideline from the Society of Vascular and Interventional Neurology Guidelines and Practice Standards Committee. Stroke Vasc Interv Neurol. 2025;5(6):e001814. doi:10.1161/SVIN.125.001814.

20. Tanaka T, Fujimoto S, Saitoh K, Satoh S, Nagamatsu K, Midorikawa H. Superselective angiographic findings of ipsilateral middle meningeal artery of chronic subdural hematoma in adults. No Shinkei Geka. 1998;26(4):339-347.

21. Torihashi K, Sadamasa N, Yoshida K, Narumi O, Chin M, Yamagata S. Independent predictors for recurrence of chronic subdural hematoma: a review of 343 consecutive surgical cases. Neurosurgery. 2008;63(6):1125–1129. doi:10.1227/01.NEU.0000335782.60059.17.

22. Shen J, Gao Y, Li Q, et al. Risk factors predicting recurrence of bilateral chronic subdural hematomas after initial bilateral evacuation. World Neurosurg. 2019;130:e133–e139. doi:10.1016/j.wneu.2019.06.016.

23. Chaliparambil RK, et al. Bilateral middle meningeal artery embolization for the treatment of bilateral chronic subdural hematoma. Clin Neurol Neurosurg. 2025;248:108664. doi:10.1016/j.clineuro.2024.108664.

