## Supplementary Results for "Burr-Hole Intersection of Middle Meningeal Artery Branches and Recurrence in Chronic Subdural Haematoma: a Multicentre Retrospective Cohort Study"

| Outcome | Variable | Adjusted OR | 95% CI | p value |
|---|---|---|---|---|
| **Radiological recurrence** | No branch hit | Reference | — | — |
| | Distal-only hit | 0.52 | 0.19–1.39 | 0.191 |
| | Main-branch hit | **0.30** | **0.11–0.81** | **0.018** |
| | Age, per 10 years | 0.91 | 0.67–1.24 | 0.560 |
| | Female sex | 0.84 | 0.36–1.95 | 0.684 |
| | Site, RLH vs NHNN | 0.67 | 0.22–2.08 | 0.488 |
| | Maximum thickness, per 5 mm | 0.90 | 0.65–1.25 | 0.529 |
| | Prior anticoagulant or antiplatelet use | 0.69 | 0.27–1.75 | 0.434 |
| **Parameters.** Patients / hemispheres = 136, events = 37, Log-likelihood = -75, AIC = 165.99, AUC = 0.671 | | | | |
| **Interventional recurrence** | No branch hit | Reference | — | — |
| | Distal-only hit | 0.69 | 0.21–2.30 | 0.546 |
| | Main-branch hit | 0.62 | 0.20–1.90 | 0.402 |
| | Age, per 10 years | 0.89 | 0.62–1.27 | 0.516 |
| | Female sex | 0.74 | 0.27–2.07 | 0.572 |
| | Site, RLH vs NHNN | 0.79 | 0.20–3.12 | 0.741 |
| | Maximum thickness, per 5 mm | 1.03 | 0.70–1.52 | 0.883 |
| | Prior anticoagulant or antiplatelet use | 0.46 | 0.13–1.54 | 0.207 |
| **Parameters.** Patients / hemispheres = 137, events = 22, Log-likelihood = -58, AIC = 132.04, AUC = 0.626 | | | | |

**Supplementary Table 1.** Patient-clustered multivariable logistic regression of branch-hit group and radiological and interventional recurrence for unilateral cSDH cases only. The intercept was included in the model but omitted from the table for brevity.

| Outcome | Variable | Adjusted OR | 95% CI | p value |
|---|---|---|---|---|
| **Radiological recurrence** | Nearest-groove distance, per 5 mm | 1.45 | **1.03–2.05** | **0.032** |
| | Age, per 10 years | 0.89 | 0.65–1.20 | 0.445 |
| | Female sex | 0.84 | 0.36–1.94 | 0.686 |
| | Site, RLH vs NHNN | 0.60 | 0.19–1.88 | 0.392 |
| | Maximum thickness, per 5 mm | 0.93 | 0.67–1.28 | 0.679 |
| | Prior anticoagulant or antiplatelet use | 0.70 | 0.28–1.76 | 0.451 |
| **Parameters.** Patients / hemispheres = 136, events = 37, Log-likelihood = -75.77, AIC = 165.54, AUC = 0.65 | | | | |
| **Interventional recurrence** | Nearest-groove distance, per 5 mm | **1.52** | **1.05–2.20** | **0.027** |
| | Age, per 10 years | 0.87 | 0.61–1.24 | 0.435 |
| | Female sex | 0.76 | 0.27–2.14 | 0.597 |
| | Site, RLH vs NHNN | 0.64 | 0.16–2.64 | 0.540 |
| | Maximum thickness, per 5 mm | 0.99 | 0.66–1.47 | 0.951 |
| | Prior anticoagulant or antiplatelet use | 0.49 | 0.14–1.66 | 0.253 |
| **Parameters.** Patients / hemispheres = 137, events = 22, Log-likelihood = -56, AIC = 126.12, AUC = 0.668 | | | | |

**Supplementary Table 2.** Patient-clustered multivariable logistic regression of nearest-groove distance and radiological and interventional recurrence for unilateral cSDH cases only. The intercept was included in the model but omitted from the table for brevity.

**Power calculation:**

Using recurrence requiring intervention as the outcome, the observed adjusted OR of 1.33 per 5-mm increase in nearest-groove distance was used for prospective observational sample-size estimation. Using the method of Hsieh et al., approximately 617 analysable hemispheres would be required to achieve 80% power at a two-sided $\alpha$ of 0.05. Allowing for the current complete-case rate of 87.3%, this corresponds to approximately 700 hemispheres[1].

For a future randomised trial, using the method of Fleiss et al. and assuming an intervention-requiring recurrence rate of 16% in the standard-care group, approximately 986 patients would be required to detect a reduction to 10% with 80% power and a two-sided $\alpha$ of 0.05, increasing to approximately 1,040 after allowing for 5% loss to follow-up[2].

Supplementary data references:


1. Hsieh FY, Bloch DA, Larsen MD. A simple method of sample size calculation for linear and logistic regression. *Stat Med.* 1998;17(14):1623-1634. doi:10.1002/(SICI)1097-0258(19980730)17:14<1623::AID-SIM871>3.0.CO;2-S.

2. Fleiss JL, Tytun A, Ury HK. A simple approximation for calculating sample sizes for comparing independent proportions. *Biometrics.* 1980;36(2):343-346. doi:10.2307/2529990.
